# Role of Weight Loss Induced Prediabetes Remission in the Prevention of Type 2 Diabetes – Time to Improve Diabetes Prevention

**DOI:** 10.1101/2024.03.22.24304576

**Authors:** Reiner Jumpertz von Schwartzenberg, Elsa Vazquez Arreola, Arvid Sandforth, Robert L. Hanson, Andreas L. Birkenfeld

## Abstract

**Aims/hypothesis:** For individuals with prediabetes, the current American Diabetes Association (ADA) guidelines recommend a body weight loss >7% to prevent the development of type 2 diabetes (T2D) without any glycemic target goals. However, we have recently shown that weight loss induced prediabetes remission reduces relative T2D risk by 73% within the following two years. Therefore, we investigated relative T2D risk reduction in people with weight loss vs those with weight loss and prediabetes remission in an independent cohort: the randomized controlled Diabetes Prevention Program (DPP).

**Methods:** Individuals who lost >7% of their body weight over the first year were included in this analysis. Of these, 416 were assigned to intensive lifestyle intervention and 64 received placebo. Remission of prediabetes was defined by normalization of fasting and 2h glucose during an oral glucose tolerance test and a normalized HbA1c according to ADA criteria. Non-remission was given when at least one of these criteria was not met. We computed Kaplan-Meier curves and compared them using log-rank tests and future T2D risk was assessed by computing the relative risk between groups.

**Results:** In DPP, 480 individuals achieved a weight loss of >7% and of these 114 additionally achieved prediabetes remission. Over the period of 6 years, those who achieved weight loss and remission had a 66% lower relative risk to develop T2D compared to those who only met the 7% weight loss goal [RR=0.34, 95% CI (0.15, 0.76)]. Similarly, weight loss responders had a lower relative future T2D risk compared to weight loss non-responders [RR=0.28, 95% CI (0.13, 0.64)]. Importantly, there was only a single T2D case in weight loss responders for up to 4 years after the intervention.

**Conclusions/interpretation:** The combination of achieving weight loss goals and prediabetes remission is most effective in reducing future T2D risk. Thus, beside weight loss goals, interventions in individuals with prediabetes should be continued until prediabetes remission is achieved and this ought to be adapted in current clinical praxis guidelines.

## Research Letter

By 2050, more than 1.3 billion people worldwide are expected to have diabetes; the vast majority of patients will have type 2 diabetes (T2D) [1]. T2D increases the risks of many chronic diseases, including micro- and macrovascular diseases, neurodegenerative disease, and cancer, posing a huge burden on affected people and societies. Recent data show that more than 80% of patients with T2D will live in low and middle-income countries and thus, T2D is becoming more and more a disease of inequity. [1]. Prevention is the only strategy to reduce future incidence of T2D and is therefore, an urgent clinical need.

People with prediabetes have a lifetime risk to develop T2D of 74% and prediabetes predisposes to diseases other than diabetes, particularly microvascular - and cardiovascular disease [2,3]. We have recently shown in a predefined post hoc analysis from the Prediabetes Lifestyle Intervention Study (PLIS) [4] with validation data from the US Diabetes Prevention Program (DPP) [4-6] that lifestyle (including dietary counseling and increased physical exercise) -induced weight loss of > 5 % led to remission of prediabetes in 73% of patients [4]. Remission rates increased with increasing weight loss. Thus, weight loss is an important driver of prediabetes remission [4] and in PLIS, insulin sensitivity was critical [4]. Previous analysis from DDP showed that younger age and insulin secretion at baseline were predictive for remission [7].

In our analysis in PLIS and DPP [4], we defined remission as return to normal glucose regulation, including all features of normal fasting glucose (<5·6 mmol/l), normal glucose tolerance (2-h post-load glucose <7·8 mmol/l) and HbA1c levels below 39 mmol/mol at the end of the lifestyle intervention. Importantly, weight loss induced prediabetes remission in PLIS not only reduced the relative risk of developing T2D, it was also associated with lower renal albumin excretion and higher skin small vessel density as assessed by raster-scanning optoacoustic mesoscopy (RSOM) [4], suggesting improved small vessel integrity.

Current American Diabetes Association (ADA) Standards of Care for prevention and delay of diabetes recommend that persons with prediabetes should lose body weight of ≥ 7%. Specific glucose target values are not recommended [8]. The 7% weight loss goal was chosen because it was feasible to achieve and maintain and was likely to reduce the risk of developing diabetes [8].

Here, we suggest that body weight loss and glycemic remission goals should be considered together, since weight loss and remission of prediabetes together offer the most effective protection against the development of T2D. To support this notion, we used data from the DPP (ClinicalTrials.gov registration no. NCT00004992) repository, which is the basis of the current Standards of Care of prevention or delay of diabetes by the ADA [8]. DPP was a randomized multicenter clinical trial that studied the effects of an intensive lifestyle intervention (ILS) and metformin on the prevention or delay of T2D in people with prediabetes. Inclusion criteria were age≥25 years, BMI≥24 kg/m2 (≥22 kg/m2 if Asian), and a diagnosis of both impaired fasting glucose and impaired glucose tolerance. Participants were recruited between July 31, 1996 and May 18, 1999 from 27 clinical centers in the USA; they were randomly assigned to receive either an ILS, metformin, or a placebo. Participants in the ILS group received 16 one-to-one lessons covering diet, exercise, and behavior modification during the first 24 weeks, with advice to engage in moderate physical activity for at least 150 minutes per week. Diabetes was diagnosed using an annual oral glucose tolerance test (OGTT) or a semiannual FPG test according to the American Diabetes Association 1997 criteria; diagnosis confirmation was done by a repeat test [9].

In DPP, we compared incident T2D over 4 and 6 years in all persons with prediabetes who reached the guideline goal of at least 7% body weight loss during the lifestyle intervention but did not go into remission (non-responders) to those who lost at least 7% of body weight and additionally reached normal glucose regulation (responders) as defined above. This secondary analysis included 480 participants randomized to the intensive lifestyle intervention (ILS) or placebo that lost at least 7% of their baseline weight by year 1, had complete measurements of HbA1c, FPG, and 2hr-PG at baseline and year 1, and had follow-up data on diabetes diagnosis; participants assigned to metformin were not included. Descriptive statistics for participants at baseline were calculated using frequency distributions for categorical variables. Continuous variables were summarized using arithmetic means and standard deviations or medians and interquartile ranges and were compared using t-tests or Wilcoxon tests as appropriate. Fisher’s exact tests were used to compare proportions of progressors and non-progressors between groups and logistic regression models were utilized to determine if the probability of progression to diabetes differed between these groups when adjusting for treatment arm. Risk of progression to T2D within the first six years of follow-up between groups was compared through relative risks adjusted for treatment. Risk of incident T2D in all groups was visualized using Kaplan Meir (KM) curves for survival probability that were adjusted for treatment. KM curves were compared using log-rank tests.

In the DPP repository data, there were 480 participants treated with ILS who lost at least 7% of body weight from baseline to year 1 and who had follow-up data on diabetes diagnosis. Descriptive characteristics of the cohort are given in the **Table**, showing that responders were younger, leaner and had lower glycemia as has been reported previously.

**Table.** Descriptive characteristics of DPP participants at baseline and 1 year.

|  | Baseline |  |  | 12-month |  |  |
| --- | --- | --- | --- | --- | --- | --- |
|  | Responders<br>(n=114) | Non-responders<br>(n=366) | p-value | Responders<br>(n=114) | Non-responders<br>(n=366) | p-value |
| Age (years), Mean (SD) | 49.8 (10.7) | 53.2 (11.4) | 0.005 |  |  |  |
| BMI (kg/m), Median (IQR) | 33.0 (29.3, 38.4) | 32.2 (28.9, 36.4) | 0.101 | 28.0 (25.6, 32.8) | 28.6 (25.4, 32.3) | 0.604 |
| Weight (kg), Median (IQR) | 94.3 (81.6, 108.8) | 89.9 (78.0, 103.0) | 0.027 | 80.3 (69.4, 95.8) | 79.0 (69.2, 91.5) | 0.279 |
| Fasting glucose (mmol/L), Median (IQR) | 5.7 (5.5, 5.9) | 5.8 (5.6, 6.2) | <0.001 | 5.2 (5.0, 5.3) | 5.6 (5.3, 5.8) | <0.001 |
| 30 min OGTT glucose (mmol/L), Median (IQR) | 9.0 (8.3, 9.9) | 9.4 (8.6, 10.4) | 0.003 | 7.8 (6.9, 9.0) | 8.8 (7.9, 9.8) | <0.001 |
| 120 min OGTT glucose (mmol/L), Median (IQR) | 8.7 (8.1, 9.7) | 9.0 (8.4, 9.9) | 0.005 | 6.0 (5.0, 6.7) | 7.8 (6.5, 9.0) | <0.001 |
| HbA1c (%), Median (IQR) | 5.5 (5.3, 5.7) | 5.9 (5.7, 6.1) | <0.001 | 5.4 (5.1, 5.6) | 5.8 (5.5, 6.0) | <0.001 |
| HbA1c (mmol/mol), Median (IQR) | 36.6 (34.4, 38.8) | 41.0 (38.8, 43.2) | <0.001 | 35.5 (32.2, 37.7) | 39.9 (36.6, 42.1) | <0.001 |
| Fasting insulin (uU/mL), Median (IQR) | 21.5 (16.0, 32.0) | 24.0 (16.0, 33.0) | 0.654 | 13.0 (9.0, 20.0) | 15.0 (11.0, 21.0) | 0.042 |
| 30 min OGTT insulin (uU/mL), Median (IQR) | 86.5 (52.5, 128.5) | 84.0 (61.0, 122.0) | 0.913 | 67.0 (41.0, 99.0) | 68.0 (45.0, 94.0) | 0.974 |
| HOMA-B, Median (IQR) | 191.6 (144.0, 290.0) | 194.6 (140.0, 266.1) | 0.431 | 170.0 (108.0, 250.9) | 150.7 (107.0, 204.3) | 0.064 |
| HOMA-IR, Median (IQR) | 5.5 (4.2, 8.1) | 6.1 (4.2, 8.8) | 0.367 | 2.9 (2.0, 4.7) | 3.8 (2.6, 5.2) | 0.001 |
| Fasting insulin sensitivity index, Median (IQR) | 0.2 (0.1, 0.2) | 0.2 (0.1, 0.2) | 0.367 | 0.3 (0.2, 0.5) | 0.3 (0.2, 0.4) | 0.001 |
| Proinsulin (pM), Median (IQR) | 12.0 (8.0, 19.0) | 14.0 (9.9, 21.0) | 0.060 | 7.0 (4.0, 11.0) | 8.3 (5.2, 13.0) | 0.015 |
| 30 min corrected insulin response, Median (IQR) | 0.6 (0.4, 0.8) | 0.5 (0.3, 0.7) | 0.052 | 0.6 (0.4, 1.0) | 0.5 (0.3, 0.7) | 0.001 |

We compared non-responders to responders among participants who lost >7% of body weight in DPP. From the 480 DPP participants who lost at least 7% of weight from baseline to year 1, 114 were responders (they remitted to normal glucose regulation at year 1) and 366 were non-responders (they did not remit to normal glucose regulation at year 1). Of the 366 non-responders, 42 were diagnosed with diabetes by year 4 of follow-up. Of the 114 responders, one was diagnosed with diabetes by year 4 of follow-up. Responders had a significantly lower adjusted relative risk of progression to diabetes compared to non-responders over the period of 6 years [RR=0.28, 95% CI (0.13, 0.64)]. By year 6 of follow-up, the proportions of people with incident T2D in responders and non-responders were markedly lower in responders (p=0.0002, Fischer’s Exact text). After adjusting for treatment arm, responders had still a significantly lower probability of progressing to diabetes compared to non-responders [OR=0.24, 95% CI (0.10, 0.58)], (p=0.0005).

Below, we show the Kaplan Meier curve for survival probability for responders and non-responders adjusted for treatment. It can be observed that after year 2, responders started to have a lower probability for diabetes development than non-responders (p=0.0005).

**Figure.**
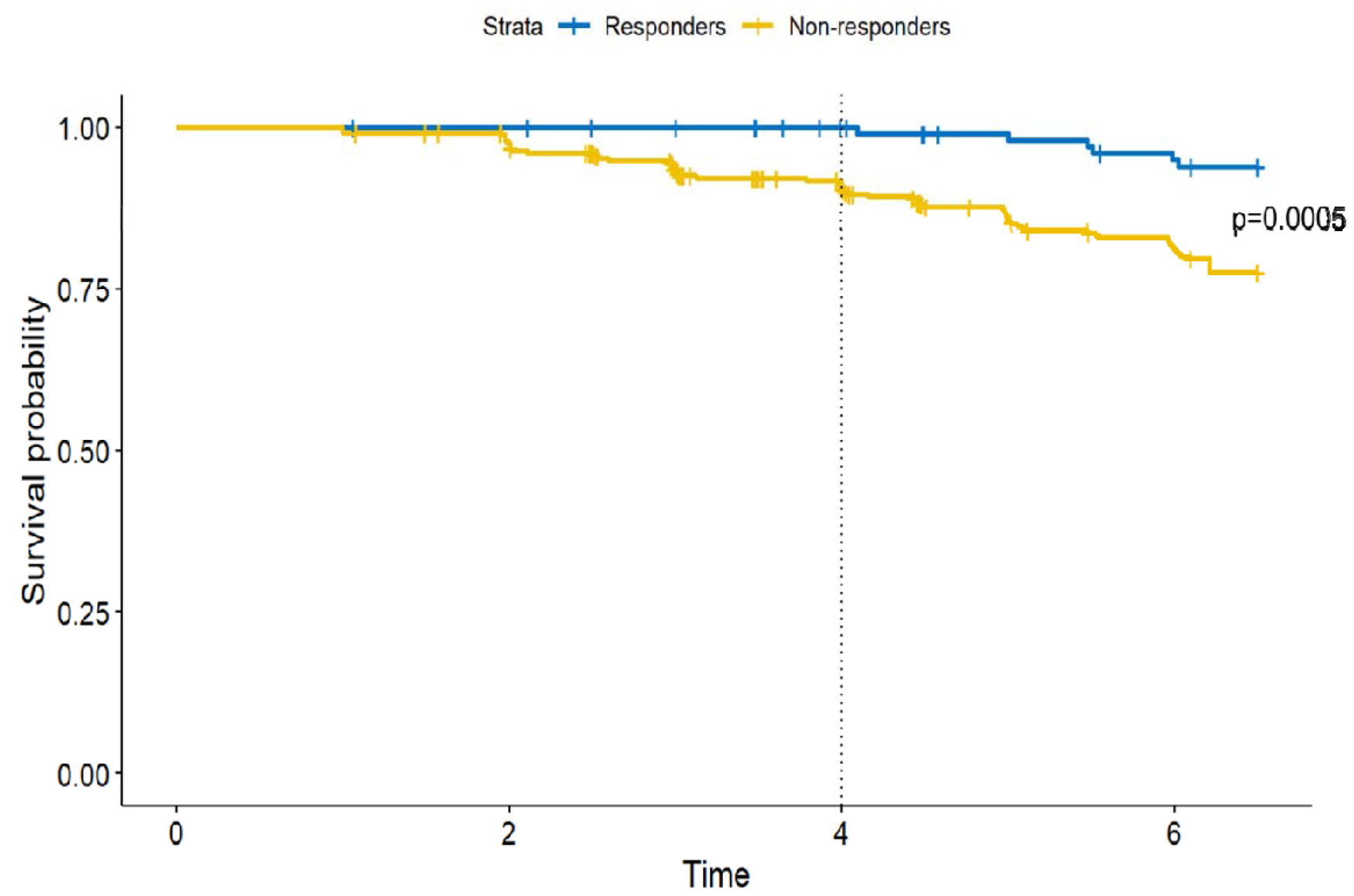
Kaplan-Meier curve for the probability of developing incident diabetes in responders and non-responders adjusted for treatment arm. After year 2, responders started to have a lower probability of developing diabetes than non-responders, leading to a continuous and progressive advantage for responders, i.e. people who achieved remission after year 1, over time (p=0.0005).

In summary, combining the recent ADA ‘Standards of Care for the Delay and Drevention of Diabetes’ recommendation for people with prediabetes to lose 7% of body weight [8] with the goal to reach remission from prediabetes to normal glucose regulation, reduces the relative risk to develop T2D by 76% within 6 years and importantly within the first 4 years, there was only one single incident diabetes case in this group, indicating that less than 1% of these patients develops diabetes after 4 years. As weight loss is a determining factor for the remission of prediabetes [6], we hypothesize that individuals with prediabetes who did not achieve remission (non-responders) after losing ≥7% of their body weight may benefit from continued weight loss until they reach their personal threshold [10,11]. Other strategies such as increasing physical exercise are also potential options [12]. We have shown previously in PLIS that more intensive lifestyle intervention is more effective in bringing patients in remission than conventional lifestyle interventions [6]. Thus, adding glycemic targets (i.e. normal glucose regulation) to weight loss goals in patients with prediabetes offers a clear, measurable and reliable goal that can be reached and that is more effective to prevent T2D than current recommendations. We advocate that preventive strategies for diabetes should include a return to normal glucose regulation, i.e. remission.

We conclude that remission of prediabetes is a critical therapeutic strategy to reduce the enormous burden of T2D in the future, especially considering the increasing global inequity of T2D. The concept of remission of prediabetes needs to be considered for future guidelines as it has the potential to reduce the increasing incidence and prevalence of T2D worldwide.

## Data Availability

In general data of the DPP cohort can be requested from the Diabetes Prevention Program website.

## Acknowledgements

ALB, RJvS, AS are grateful to the support by the German Federal Ministry for Education and Research (01GI0925) via the German Center for Diabetes Research (DZD e.V.); Ministry of Science, Research and the Arts Baden-Württemberg and Helmholtz Munich.

The Diabetes Prevention Program (DPP) was conducted by the DPP Research Group and supported by the National Institute of Diabetes and Digestive and Kidney Diseases (NIDDK), the General Clinical Research Center Program, the National Institute of Child Health and Human Development (NICHD), the National Institute of Aging (NIA), the Office of Research on Women’s Health, the Office of Research and Minority Health, the Centers for Disease Control and Prevention (CDC), and the American Diabetes Association. The data (and samples) from the DPP were supplied by the NIDDK Central Repository. This manuscript was not prepared under the auspices of the DPP and does not represent analyses or conclusion of the DPP Research Group, the NIDDK Central Repository, or the NIH.

## Funding

The analysis for this paper was supported by the Intramural Research Program of the National Institute of Diabetes and Digestive and Kidney Diseases (NIDDK). DPP was supported by NIDDK and other agencies. German Federal Ministry for Education and Research (01GI0925) via the German Center for Diabetes Research (DZD e.V.); Ministry of Science, Research and the Arts Baden-Württemberg; Helmholtz Munich. ALB was supported by a research grant from the German Research Foundation (GRK2816; BI 1292/9-1; BI 1292/10-1; BI1292/ 12-1). RJvS was funded by Helmholtz Munich and the Helmholtz Association via a Helmholtz Young Investigator Group (VH-NG-1619) and by the Cluster of Excellence (EXC-2124/1-03.007).

The study funder was not involved in the design of the study; the collection, analysis, and interpretation of data; writing the report; and did not impose any restrictions regarding the publication of the report.

## Authors’ relationships and activities

ALB received lecture / advisory board fees from Bayer; lecture fees from NovoNordisk, Boehringer Ingelheim, Daiichi Sankyo, Lilly paid to University Hospital Tübingen: ALB is co-founder of Eternygen GmbH

## Contribution statement

ALB is the guarantor for the work and conceptualized the details of this analysis. RJvS, EVA, AS and RLH accessed and analyzed the data. ALB wrote the manuscript; RJvS, EVA, RLH and AS edited the manuscript.

## Notes

### Author Declarations

The study used data of DPP which can be requested here: https://repository.niddk.nih.gov/studies/dpp/

### Summary of Updates

As there was new scientific evidence, we corrected one statement in the abstract

